# Temporal trends and factors associated with emergency department visits and hospitalizations in angina with no obstructive coronary artery disease (ANOCA)

**DOI:** 10.1101/2023.07.20.23292978

**Authors:** Shubh Patel, Marinda Fung, Sonia Butalia, Todd J Anderson

## Abstract

**Introduction:** Angina with no obstructive coronary artery disease (ANOCA) presents diagnostic and treatment challenges, significantly burdening healthcare resources as reported in previous studies. This study assessed temporal changes in emergency department (ED) visits and hospitalizations for ANOCA and factors associated with these outcomes.

**Methods/Results:** We assessed a retrospective cohort of 85,573 individuals (26% with ANOCA, 31% female, mean age 62.1±12.0 years) who underwent their first cardiac catheterization for chest pain in Alberta from 2002 to 2017. Temporal trend analysis showed ED visits ranged from 26.3% to 30.7% (β=0.21 [95%CI:-0.28 to 0.70];p=0.33) for ANOCA and from 47.7% to 53.1% (β=-0.15[95%CI:-0.87 to 0.57];p=0.63) for obstructive CAD, with no temporal changes in either. Hospitalizations decreased from 6.5% to 3.8% (β=-0.28 [95%CI:-0.47 to -0.09];p=0.010) for ANOCA and from 24.8% to 15.3% (β=-1.45 [95%CI:-1.77 to -1.12];p<0.001) for obstructive CAD. Multivariable logistic regression analysis factors associated with ED visits in individuals with ANOCA included cerebrovascular disease (CEVD) (OR=1.73 [95%CI:1.40–2.15]), congestive heart failure (CHF) (OR=1.91 [95%CI:1.49–2.44]), peripheral artery disease (PAD) (OR=1.61 [95%CI:1.18–2.19), and unstable angina (UA) (versus (vs) stable angina (SA): OR=1.65 [95%CI:1.51–1.80]). Factors associated with hospitalizations in ANOCA included CEVD (OR=1.39 [95%CI:1.11–1.73]), CHF (OR=2.06 [95%CI:1.66–2.56]), hypertension (OR: 1.26 [95%CI:1.14–1.40]), PAD (OR=1.89 [95%CI:1.43–2.50]), and myocardial infarction (vs SA: OR=1.27 [95%CI:1.12–1.44]), and UA (vs SA: OR=1.36 [95%CI:1.22–1.52]).

**Conclusions:** ED visits for ANOCA remained stable, while hospitalizations declined over time. Understanding factors associated with recurrent visits may aid clinicians in treatment strategies.

**Clinical Perspective:** *What is new?:*

- This study was the first to characterize emergency department (ED) visits and assess temporal trends of ED visits and hospitalizations in individuals with ANOCA.
- ED visits for individuals with ANOCA or obstructive CAD have remained stable over time, with a significant decline in hospitalizations for both groups.
- In contrast to previous studies, individuals with obstructive CAD had higher ED visits and hospitalizations rates than those with ANOCA.

*What are the clinical implications?:*

- Increased recognition of ANOCA may be responsible for decreasing hospitalizations.
- Despite increased awareness, individuals with ANOCA still frequently visit the ED, highlighting the need for improved education on persistent chest pain and unnecessary ED visits.
- The development of specialized chest pain clinics using a multidisciplinary approach could reduce unnecessary ED visits and hospitalizations, thereby improving the quality of life for individuals with ANOCA.

## Introduction

Chest pain is a common cause of emergency department (ED) visits and hospitalizations.^1, 2^ While most individuals presenting with chest pain to the ED do not have a cardiac etiology, around 20% are diagnosed with coronary artery disease (CAD) as the cause of their symptoms.^3^ For those with ischemic causes, a significant proportion have angina with no obstructive coronary artery disease (ANOCA). This subgroup of individuals poses a challenge in diagnosis and treatment, leading to considerable healthcare resource use, such as hospitalizations and repeat cardiac catheterizations.^4–11^ ANOCA has been associated with poor quality of life, increased disability risk, and higher rates of adverse events.^10, 12–15^

Despite the growing recognition of ANOCA and its link to recurrent chest pain, the temporal trends in ED visits and hospitalizations for individuals with ANOCA remain unclear.^16^ To our knowledge, no studies have assessed the trends of ED visits and hospitalizations in ANOCA. However, numerous studies have underscored that individuals diagnosed with ANOCA significantly impact healthcare resource use, primarily due to frequent hospitalizations.^5, 7, 8, 10^ These studies tend to be limited by their inclusion of highly selective and symptomatic individuals, and considering the rapidly growing acknowledgement of ANOCA, they could be deemed outdated. Such recognition of this entity could have altered hospitalization trends over time. Consequently, our objective was to assess changes in ED visits and hospitalizations for ANOCA over time and identify the factors associated with these outcomes.

We hypothesized that there had been no significant changes in the temporal trends of ED visits and hospitalizations among individuals with ANOCA. By identifying the factors associated with ED visits and hospitalizations, our research will guide the development of targeted interventions and support tools to reduce the burden of ANOCA, optimize care pathways, and improve patient outcomes.

## Methods

### Ethics

Following the University of Calgary’s Institutional Ethics Review Board’s approval, we conducted a retrospective study with informed participant consent at the time of cardiac catheterization. We pooled anonymous data analysis authorized by a consent waiver.

### Study Population

The study population was extracted from the Alberta Provincial Project for Outcome Assessment in Coronary Heart Disease (APPROACH) database, comprising adults (≥18 years) in Alberta who underwent initial cardiac catheterization due to chest pain since 1995.^17^ The database, enriched with administrative records, offers comprehensive clinical and cardiovascular treatment details.^18^ We included individuals whose first cardiac catheterization fell between April 1, 2002, and December 31, 2017, requiring a minimum of two years of follow-up data and a valid Personal Health Number (PHN) for data linkage. These individuals’ ED visits and hospitalization data were tracked until March 31, 2020, for analysis. Exclusion criteria included individuals with incomplete data or unavailable data, history of coronary artery bypass grafting (CABG), percutaneous coronary intervention (PCI), left main disease, left ventricular ejection fraction (LVEF) ≤ 50%, and non-chest pain prespecified indications. Individuals with unknown LVEF were included, given that their demographic profile did not clinically differ from those with LVEF >50% and inconsistent quantification of LVEF in later years (data not shown).

Information on pre-catheterization non-invasive testing results was unavailable.

### Data Sources and Measures

Using the anonymized PHN, we linked APPROACH data to clinical and administrative databases, facilitating comprehensive data capture and linkage. These databases include the National Ambulatory Care Reporting System Database (NACRS)^19^ and the Discharge Abstract Database (DAD),^2^ providing detailed diagnostic and procedural information for all ED visits and hospitalizations in Alberta. ED visits and hospitalizations were defined as one or more incidents due to any of the prespecified ICD-10-CA codes identified in Supplemental Table S1.

Data collected from APPROACH included comorbidities, laboratory results, medications at cardiac catheterization, and sociodemographic characteristics. Coronary stenosis extent and LVEF results from cardiac catheterization were also recorded.^17^ Body mass index (BMI) was calculated as weight (kg) divided the height (m) squared, and obesity was defined as a body mass index ≥30 kg/m^2^. Renal disease was defined as an estimated glomerular filtration rate under 60mL/min/1.73 m^2^; cerebrovascular disease (CEVD) was defined as having a stroke, transient ischemic attack, or procedures for carotid artery stenosis; peripheral artery disease (PAD) was defined a history of claudication, documented obstructive peripheral vascular disease, or procedures for peripheral artery stenosis; and family history of premature coronary artery disease (FhxCAD) was defined as males <55 years of age and females <65 years presenting with CAD. Congestive heart failure (CHF) was defined as per the referring cardiologist. Smoking was defined as being a past or current smoker based on self-report.

### Study Exposures and Outcomes

ANOCA was defined as either normal coronary arteries (0% stenosis in all epicardial coronary arteries) or minimal disease (>0% but <50% stenosis in any epicardial coronary artery).

Obstructive CAD was defined as ≥ 50% stenosis in any major epicardial coronary artery. Both cohorts encompassed individuals with stable angina (SA) or acute coronary syndrome (ACS). ACS included unstable angina (UA), ST-elevation myocardial infarction (STEMI), or non-ST-elevation myocardial infarction (NSTEMI) based on the universal definition of myocardial infarction (MI).^20^

The primary outcome assessed the temporal trends in the prevalence of ANOCA and its coexistence with SA, MI, and UA among those with ED visits and hospitalizations within two years post-catheterization. The secondary outcomes were the factors associated with ED visits and hospitalizations among each group (ANOCA and obstructive CAD).

### Statistical analysis

Continuous variables were presented as mean ± standard deviation and categorical variables as percentage. Statistical comparisons between ANOCA and obstructive CAD and between individuals with and without ED visits or hospitalizations were conducted using Student’s t-test for continuous variables and the χ² test for categorical variables.

Temporal trends in the prevalence of at least one ED visit or hospitalization within two years post-catheterization among individuals with ANOCA or obstructive CAD pooled in two years intervals from 2002 to 2017 were examined using a linear regression model. Subgroup temporal trend analyses were conducted for individuals diagnosed with ANOCA categorized by presenting diagnosis—SA, MI, or UA. Beta coefficients (β), corresponding 95% confidence intervals (CIs), and p-values were reported.

Multivariable logistic regression models were developed to assess the factors associated with ED visits and hospitalizations in individuals with ANOCA and obstructive CAD. The dichotomous dependent variables were the occurrence of ED visits or hospitalizations over the study period. The process for covariate selection, detailed in Supplemental Tables S2 and S3, involved two steps: a univariable analysis followed by a multivariable analysis using the backward stepwise likelihood ratio selection procedure. While atrial fibrillation (Afib) was significant for both univariable analyses, it was excluded due to limited data availability. Odds ratios (ORs) and their corresponding 95% CIs were reported.

Statistical analyses were conducted using IBM SPSS (v29.0, IBM Corp., Armonk, NY, USA) or R statistical software (v4.2.2, R Core Team, Vienna, Austria). Significance was set at p<0.05, and all statistical tests were two-sided.

## Results

The study cohort included 85,573 individuals (26% with ANOCA, 31% female, and a mean age of 62.1 ± 12.0 years) (Figure 1; Table 1). Overall, 65% had at least one ED visit, and 30% had at least one hospitalization during the study period (Supplemental Table S4). Notably, a higher percentage of females were observed in the ANOCA group (48% vs 25%; p<0.001).

**Figure 1.**
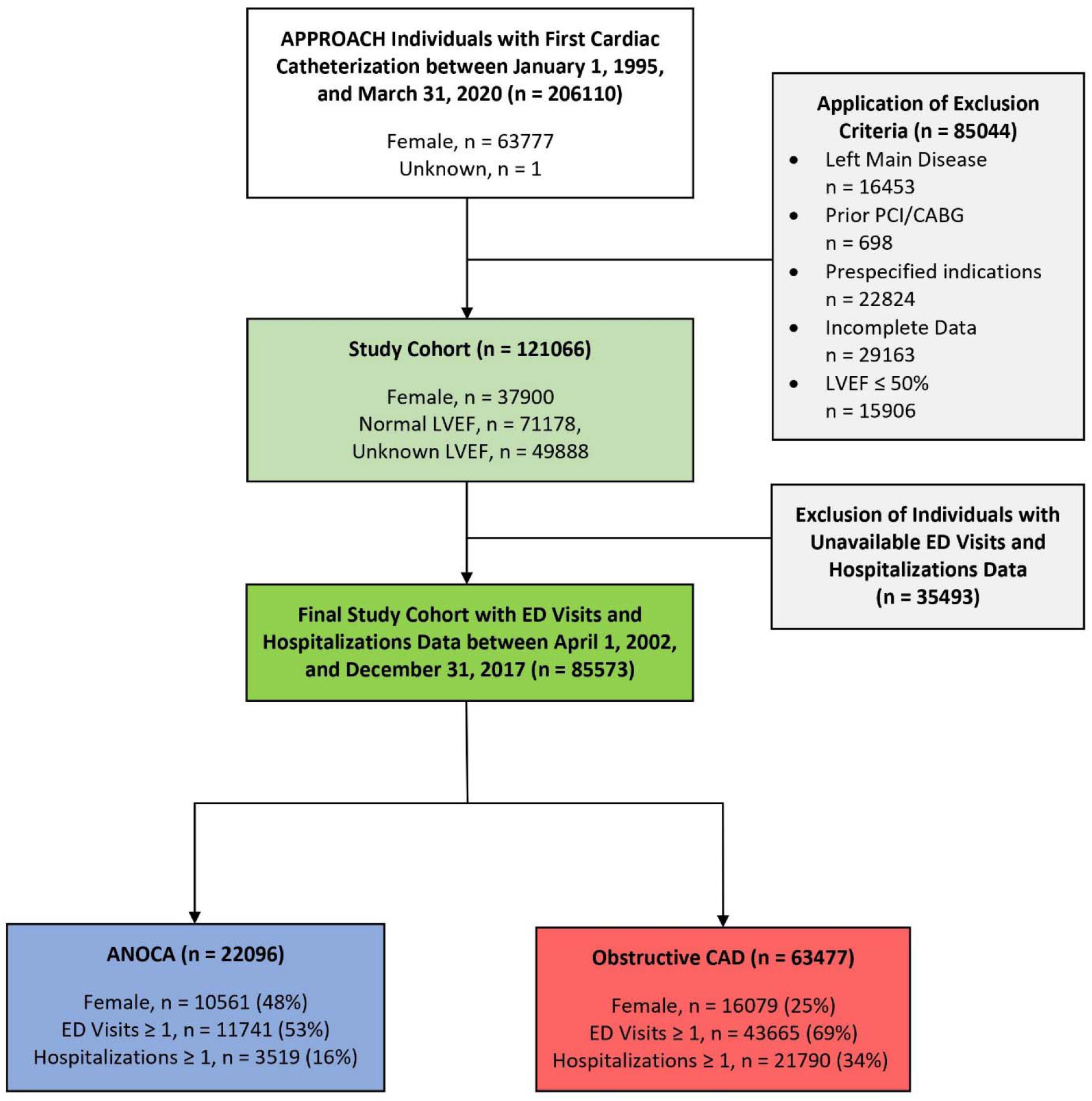
Final cohort flow. The final cohort (n = 85573) including individuals who had their first cardiac catheterization between April 1, 2002 and December 31, 2017 was derived after excluding individuals with left main disease, prior PCI/CABG, prespecified indications, incomplete data, LVEF ≤50%, and unavailable ED visits and hospitalizations data. The cohort was further divided into two subgroups: ANOCA (n = 22096) and obstructive CAD (n=63477). *APPROACH, Alberta Provincial Project for Outcome Assessment in Coronary Heart Disease; CABG, coronary artery bypass graft surgery; CAD, coronary artery disease; ED, emergency department; ANOCA, angina with no obstructive coronary artery disease; LVEF, left ventricular ejection fraction; PCI, percutaneous coronary intervention*.

**Table 1.** Baseline characteristics of individuals with ANOCA and obstructive CAD.

|  | **All**<br>**(n = 85573)** | **ANOCA**<br>**(n = 22096)** | **Obstructive CAD**<br>**(n = 63477)** | **P-value** |
| --- | --- | --- | --- | --- |
| Age, years | 62.1 ± 12.0 | 58.8 ± 12.2 | 63.3 ± 11.6 | <0.001 |
| Female | 31 | 48 | 25 | <0.001 |
| ANOCA | 26 | 100 | 0 | N/A |
| Systolic BP, mm Hg | 122.4 ± 24.7 | 123.2 ± 23.6 | 122.2 ± 25.1 | 0.001 |
| Diastolic BP, mm Hg | 68.5 ± 12.5 | 69.6 ± 11.9 | 68.2 ± 12.8 | <0.001 |
| BMI, kg/m<sup>2</sup> | 29.2 ± 6.0 | 29.7 ± 6.7 | 29.1 ± 5.7 | <0.001 |
| †ACS | 66 | 52 | 71 | <0.001 |
| MI | 73 | 52 | 79 | <0.001 |
| UA | 27 | 48 | 21 | <0.001 |
| SA | 34 | 48 | 29 | <0.001 |
| *Hospitalizations ≥ 1 | 30 | 16 | 34 | <0.001 |
| *ED visits ≥ 1 | 65 | 53 | 69 | <0.001 |
| **‡Comorbidities** |  |  |  |  |
| Hypertension | 68 | 63 | 69 | <0.001 |
| Dyslipidemia | 77 | 72 | 79 | <0.001 |
| Diabetes | 20 | 16 | 21 | <0.001 |
| Smoking | 67 | 60 | 69 | <0.001 |
| FhxCAD | 46 | 47 | 45 | 0.004 |
| Malignancy | 4 | 4 | 4 | 0.97 |
| Afib | 5 | 7 | 4 | <0.001 |
| CEVD | 5 | 4 | 6 | <0.001 |
| CHF | 4 | 4 | 4 | 0.092 |
| Renal disease | 4 | 3 | 5 | <0.001 |
| PAD | 4 | 2 | 5 | <0.001 |
| Liver disease | 1 | 1 | 1 | <0.001 |
| Obesity | 38 | 41 | 37 | <0.001 |
| **‡Medications** |  |  |  |  |
| Aspirin | 82 | 79 | 83 | <0.001 |
| Beta-blockers | 62 | 59 | 63 | <0.001 |
| Statins | 59 | 56 | 60 | <0.001 |
| CCBs | 11 | 11 | 11 | 0.098 |
| ACE-inhibitor | 43 | 38 | 45 | <0.001 |
| Long-acting nitrates | 14 | 13 | 14 | 0.002 |
| Insulin | 5 | 4 | 6 | <0.001 |
| **‡Laboratory Results** |  |  |  |  |
| TChol, mmol/L | 4.6 ± 1.2 | 4.5 ± 1.2 | 4.6 ± 1.2 | <0.001 |
| HDL-C, mmol/L | 1.2 ± 0.4 | 1.3 ± 0.4 | 1.2 ± 0.4 | <0.001 |
| LDL-C, mmol/L | 2.6 ± 1.0 | 2.5 ± 1.0 | 2.7 ± 1.0 | <0.001 |
| Triglyceride, mmol/L | 1.7 ± 1.2 | 1.6 ± 1.0 | 1.8 ± 1.3 | <0.001 |
| Random glucose, mmol/L | 7.3 ± 3.3 | 6.5 ± 2.6 | 7.6 ± 3.4 | <0.001 |
| Creatinine, µmol/L | 90.1 ± 51.1 | 85.5 ± 52.2 | 91.7 ± 50.6 | <0.001 |
| HbA1c, % | 6.4 ± 1.5 | 6.1 ± 1.4 | 6.4 ± 1.6 | <0.001 |

\*Study period from April 1, 2002 to March 31, 2020. †Some ACS patients were not subclassified; ‡Variables with incomplete data.
Data was presented in mean ± standard deviation or %. P-value assessed by Student's t-test or $\chi^2$ test, as appropriate, and significance was set at $p < 0.05$ . *ACE*, angiotensin-converting enzyme; *ACS*, acute coronary syndrome; *ANOCA*, angina with no obstructive coronary artery disease; *Afib*, atrial fibrillation; *BMI*, body mass index; *BP*, blood pressure; *CAD*, coronary artery disease; *CCB*, calcium channel blockers; *CEVD*, cerebrovascular disease; *CHF*, congestive heart failure; *ED*, emergency department; *FhxCAD*, family history of premature coronary artery; *HDL-C*, high-density lipoprotein cholesterol; *HbA1c*, hemoglobin A1c; *LDL-C*, low-density lipoprotein cholesterol; *MI*, myocardial infarction; *N/A*, not applicable; *PAD*, peripheral artery disease; *SA*, stable angina.; *TChol*, total cholesterol; *UA*, unstable angina.

### Frequency of ED visits & hospitalizations within two years post-catheterization

Individuals with ANOCA had lower ED visits and hospitalization rates than those with obstructive CAD (Table 2). Among those with ANOCA, 28.6% had at least one ED visit, and 4.8% had at least one hospitalization, whereas, in the obstructive CAD group, these frequencies were 50.0% and 19.0%, respectively. Across all categories of ED visits and hospitalizations frequencies (1-2, 3-5, and >5), individuals with ANOCA consistently showed lower proportions than those with obstructive CAD. Stable coronary diseases were the primary cause of repeat hospitalization for all individuals and those with an initial presentation of SA, MI, or UA (Supplemental Table S5).

**Table 2.**
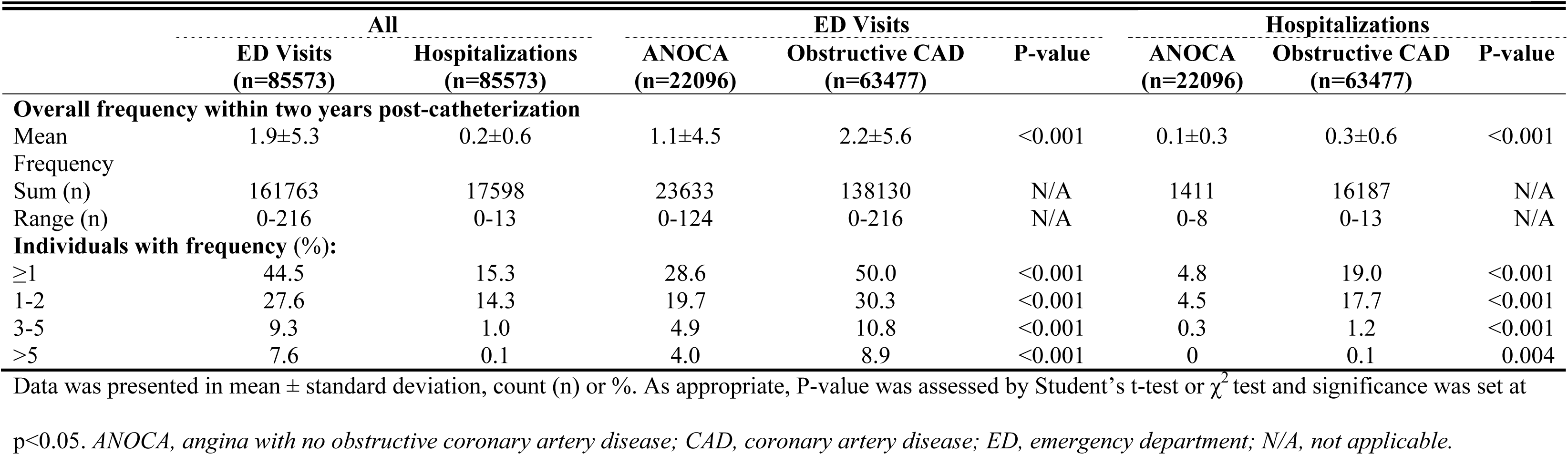
ED visits and hospitalizations frequency within two years post-catheterization.

### Temporal trends of ED visits and hospitalizations within two years post-catheterization

The prevalence of individuals with an ED visit ranged between 26.3% to 30.7% in ANOCA and 47.7% to 53.1% in obstructive CAD within two years post-catheterization over time (Figure 2A). Linear regression showed no change in ED visits for both ANOCA (β=0.21 [95% CI: **-**0.28 to 0.70]; p=0.33) and obstructive CAD (β= **-**0.15 [95% CI: -0.87 to 0.57]; p=0.63). Differences were noted in the trends depending on the initial presenting diagnosis. An increase over time was observed in individuals with ANOCA and SA (β=0.44 [95% CI: 0.17–0.71]; p=0.008) or ANOCA and UA (β=0.73 [95% CI: 0.02–1.43]; p=0.045) having an ED visit, while a reduction was noted for those with ANOCA and MI (β=**-**1.07 [95% CI: **-**1.76 to **-**0.38]; p=0.009) (Figure 2B).

**Figure 2.**
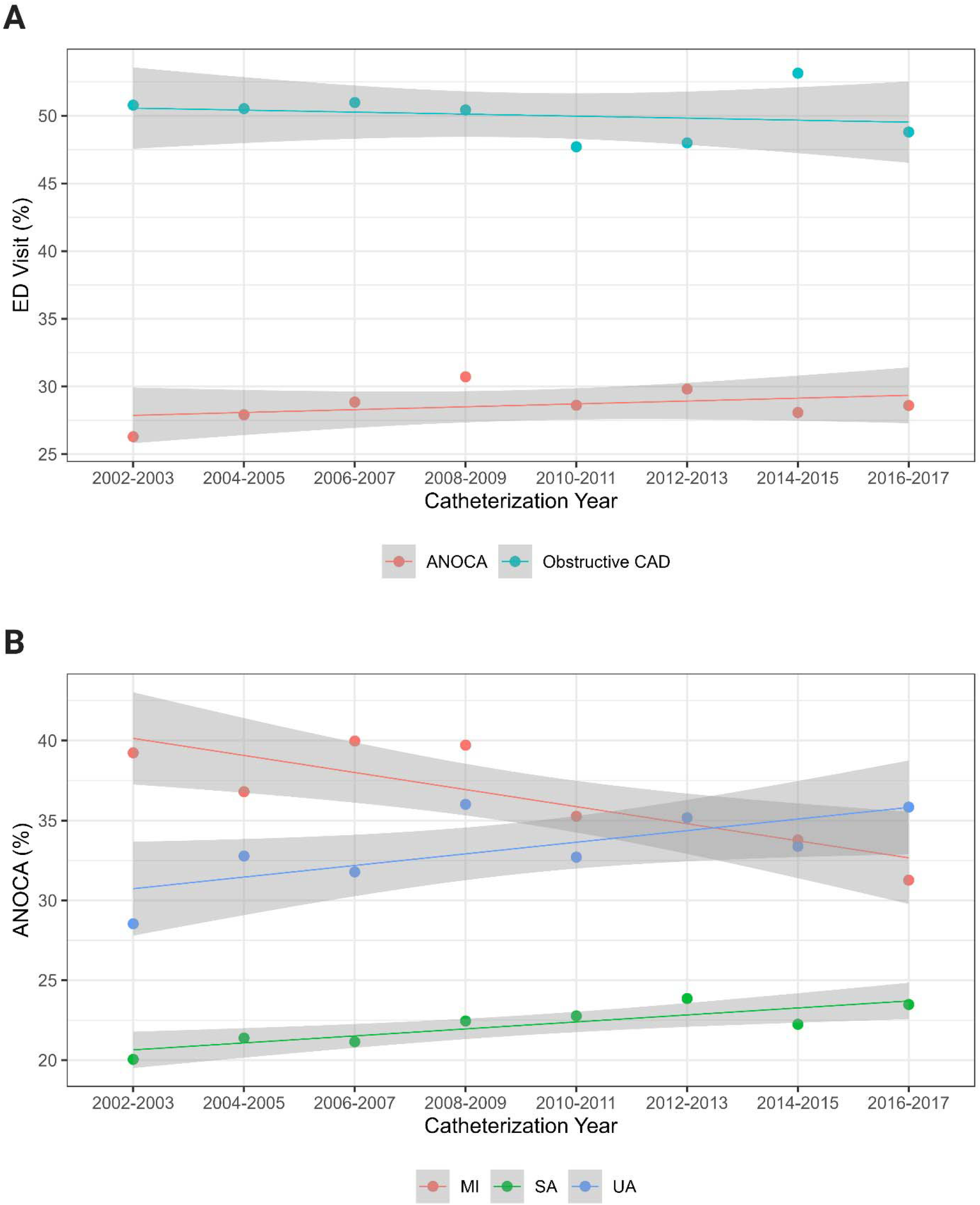
Prevalence of at least one ED visit within two years post-catheterization from 2002 to 2017. **A.** Trends in ED visits among individuals with ANOCA (β=0.21 [95% CI: **-**0.28 to 0.70]; p=0.33) and obstructive CAD (β=-0.15 [95% CI: **-**0.87 to 0.57]; p=0.63). **B.** Trends in ED visits among individuals with ANOCA and either SA (β=0.44 [95% CI: 0.17–0.71]; p=0.008), MI (β=**-**1.07 [95% CI: **-**1.76 to **-**0.38]; p=0.009), or UA (β=0.73 [95% CI: 0.02–1.43]; p = 0.045). *ANOCA, angina with no obstructive coronary artery disease. CAD, coronary artery disease; CI, confidence interval; ED, emergency department; MI, myocardial infarction; SA, stable angina; UA, unstable angina*.

The prevalence of individuals with hospitalization decreased from 6.5% to 3.8% in ANOCA and ranged between 24.8% and 15.3% in obstructive CAD (Figure 3A). Linear regression showed a decrease in hospitalizations for both ANOCA (β=-0.28 [95% CI: -0.47 to **-**0.09]; p=0.010) and obstructive CAD (β=-1.45 [95% CI: **-**1.77 to **-**1.12]; p<0.001). The prevalence of hospitalizations in individuals with ANOCA and SA, MI, or UA ranged between 2.5% to 5.2%, 5.1% to 9.8%, and 4.0% to 6.9%, respectively (Figure 3B). Similarly, individuals with ANOCA and MI (β=**-** 0.55 [95% CI: **-**0.99 to **-**0.10]; p=0.024) or ANOCA and UA (β=**-**0.34 [95% CI: **-**0.67 to **-**0.09]; p=0.043) showed a decrease in hospitalizations, whereas individuals with ANOCA and SA (β =**-** 0.19 [95% CI: **-**0.50 to 0.13]; p=0.20) had no change. The linear regression for each analysis demonstrated similar trends when stratified by sex (Supplemental Figures S1 & S2).

**Figure 3.**
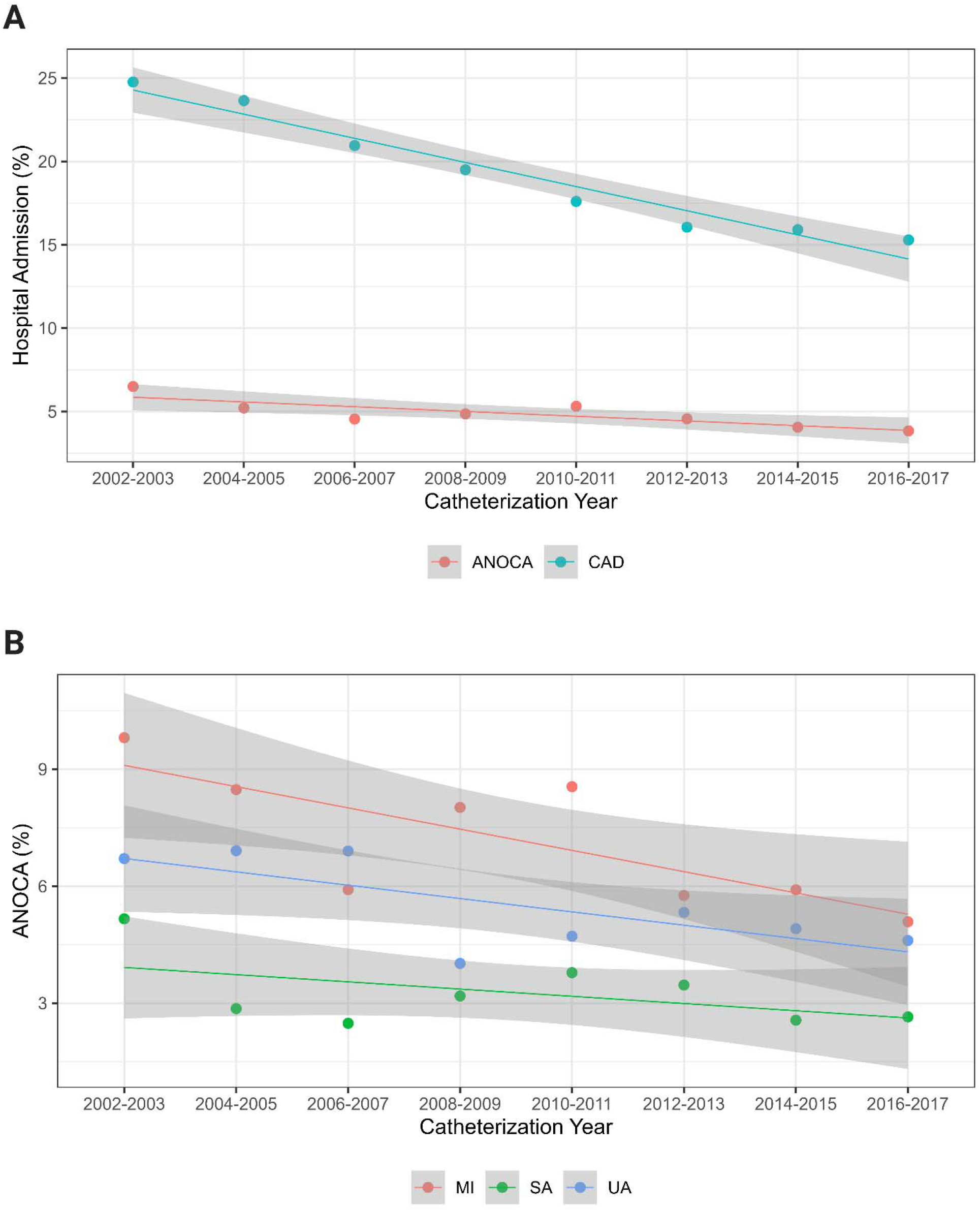
Prevalence of at least one hospitalization within two years post-catheterization from 2002 to 2017. **A.** Trends in hospitalizations among individuals with ANOCA (β= **-**0.28 [95% CI: **-**0.47 to **-**0.09]; p = 0.010) and obstructive CAD (β=**-**1.45 [95% CI: **-**1.77 to **-**1.12]; p<0.001). **B.** Trends in hospitalizations among individuals with ANOCA and either SA (β=**-**0.19 [95% CI: **-**0.50–0.13]; p =0.20), MI (β=-0.55 [95% CI: **-**0.99 to **-**0.10]; p=0.024), or UA (β=**-**0.34 [95% CI: **-**0.67 to **-**0.12]; p=0.043). *ANOCA, angina with no obstructive coronary artery disease. CAD, coronary artery disease; MI, myocardial infarction; SA, stable angina, UA, unstable angina*.

### Factors associated with ED visits and hospitalizations in ANOCA and obstructive CAD

Among individuals with ANOCA, a history of CEVD (OR=1.73 [95% CI: 1.40–2.15]), CHF (OR=1.91 [95% CI: 1.49–2.44]), PAD (OR=1.61 [95% CI: 1.18–2.19]), MI (vs SA: OR=1.41 [95% CI:1.29–1.54]), and UA (vs SA: OR=1.65 [95% CI: 1.51–1.80]) were most associated with ED visits (Table 3). For individuals with obstructive CAD, PAD (OR=1.26 [95% CI: 1.08– 1.48]), CEVD (OR=1.26 [95% CI: 1.11–1.44]), and UA (vs SA: OR=1.45 [95% CI: 1.33–1.57]) surfaced as the most significant factors.

**Table 3.**
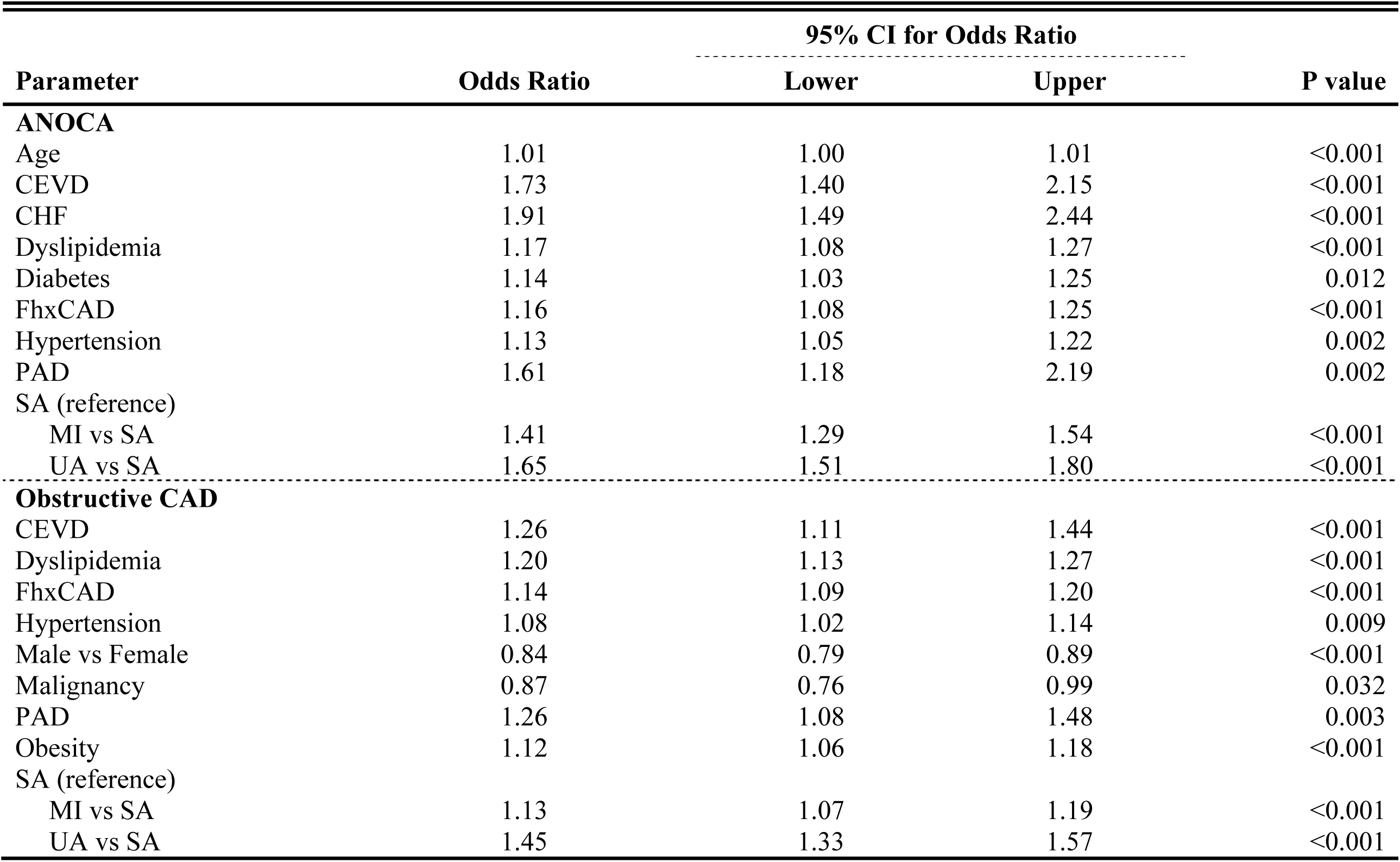
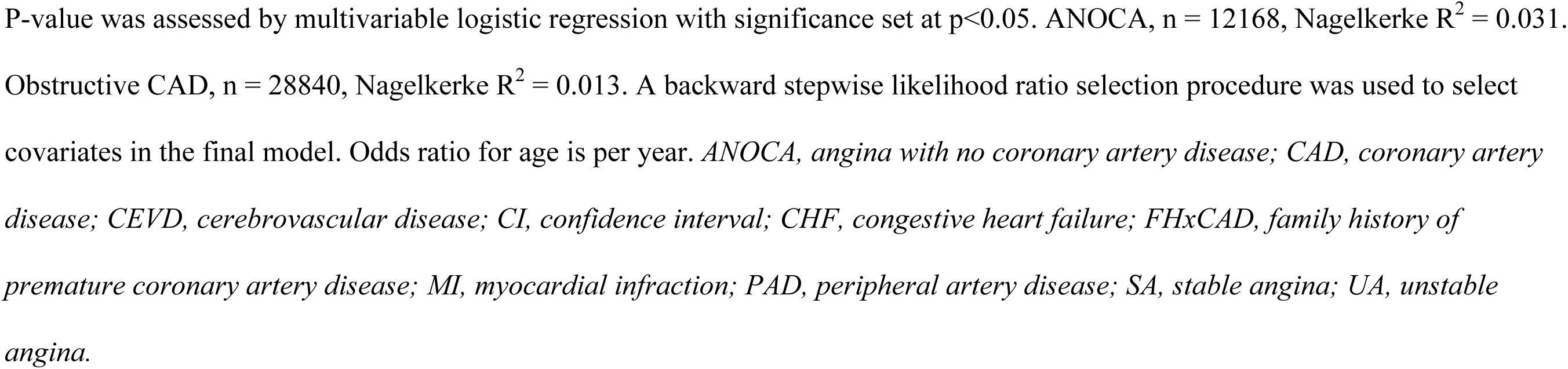
Multivariable logistic regression model defining the factors associated with ED visits in ANOCA and obstructive CAD.

| Parameter | Odds Ratio | 95% CI for Odds Ratio |  | P value |
| --- | --- | --- | --- | --- |
|  |  | Lower | Upper |  |
| ANOCA |  |  |  |  |
| Age | 1.01 | 1.00 | 1.01 | <0.001 |
| CEVD | 1.73 | 1.40 | 2.15 | <0.001 |
| CHF | 1.91 | 1.49 | 2.44 | <0.001 |
| Dyslipidemia | 1.17 | 1.08 | 1.27 | <0.001 |
| Diabetes | 1.14 | 1.03 | 1.25 | 0.012 |
| FhxCAD | 1.16 | 1.08 | 1.25 | <0.001 |
| Hypertension | 1.13 | 1.05 | 1.22 | 0.002 |
| PAD | 1.61 | 1.18 | 2.19 | 0.002 |
| SA (reference) |  |  |  |  |
| MI vs SA | 1.41 | 1.29 | 1.54 | <0.001 |
| UA vs SA | 1.65 | 1.51 | 1.80 | <0.001 |
| Obstructive CAD |  |  |  |  |
| CEVD | 1.26 | 1.11 | 1.44 | <0.001 |
| Dyslipidemia | 1.20 | 1.13 | 1.27 | <0.001 |
| FhxCAD | 1.14 | 1.09 | 1.20 | <0.001 |
| Hypertension | 1.08 | 1.02 | 1.14 | 0.009 |
| Male vs Female | 0.84 | 0.79 | 0.89 | <0.001 |
| Malignancy | 0.87 | 0.76 | 0.99 | 0.032 |
| PAD | 1.26 | 1.08 | 1.48 | 0.003 |
| Obesity | 1.12 | 1.06 | 1.18 | <0.001 |
| SA (reference) |  |  |  |  |
| MI vs SA | 1.13 | 1.07 | 1.19 | <0.001 |
| UA vs SA | 1.45 | 1.33 | 1.57 | <0.001 |

P-value was assessed by multivariable logistic regression with significance set at $p < 0.05$ . ANOCA, $n = 12168$ , Nagelkerke $R^2 = 0.031$ . Obstructive CAD, $n = 28840$ , Nagelkerke $R^2 = 0.013$ . A backward stepwise likelihood ratio selection procedure was used to select covariates in the final model. Odds ratio for age is per year. *ANOCA, angina with no coronary artery disease; CAD, coronary artery disease; CEVD, cerebrovascular disease; CI, confidence interval; CHF, congestive heart failure; FHxCAD, family history of premature coronary artery disease; MI, myocardial infarction; PAD, peripheral artery disease; SA, stable angina; UA, unstable angina.*

Salient factors associated with hospitalizations in individuals with ANOCA included CEVD (OR=1.39 [95% CI: 1.11–1.73]), CHF (OR=2.06 [95% CI: 1.66–2.56]), hypertension (OR=1.26 (95% CI: 1.14–1.40]), PAD (OR=1.89 [95% CI: 1.43–2.50]), MI (vs SA: OR=1.27 [95% CI: 1.12–1.44]), and UA (vs SA: OR=1.36 [95% CI: 1.22–1.52]) (Table 4). For those with obstructive CAD, CEVD (OR=1.34 [95% CI: 1.19–1.51]), CHF (OR=1.47 [95% CI: 1.27–1.70]), diabetes (OR=1.38 [95% CI: 1.30–1.47]), PAD (OR=1.35 [95% CI: 1.18–1.55]), and renal disease (OR=1.36 [95% CI: 1.20–1.54]) emerged as associated factors. Compared to SA, a history of either MI (OR=0.67 [95% CI 0.63–0.71]) or UA (OR=0.86 [95% CI 0.80–0.92]) was associated with a lower odds of hospitalization in individuals with obstructive CAD.

**Table 4.**
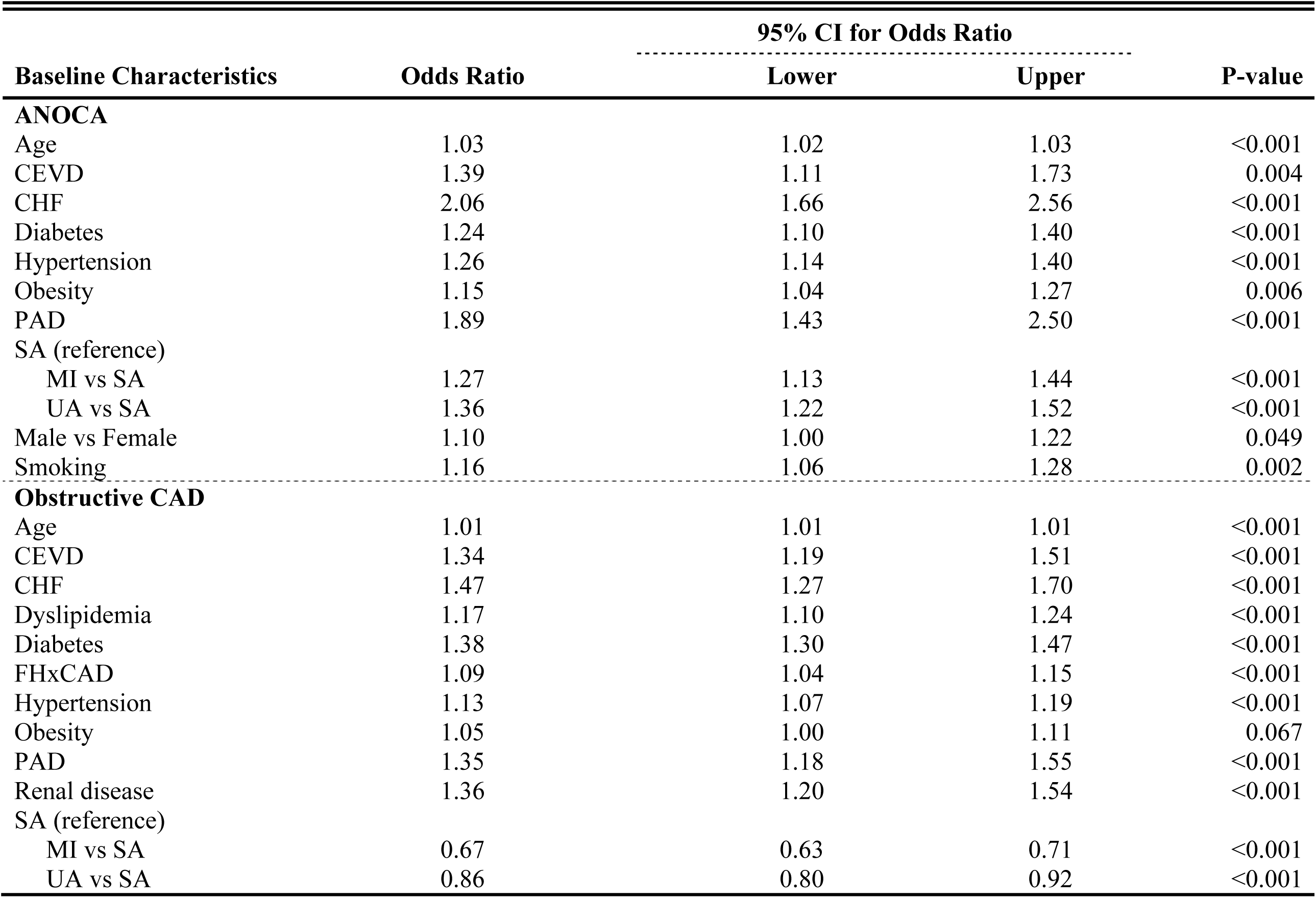
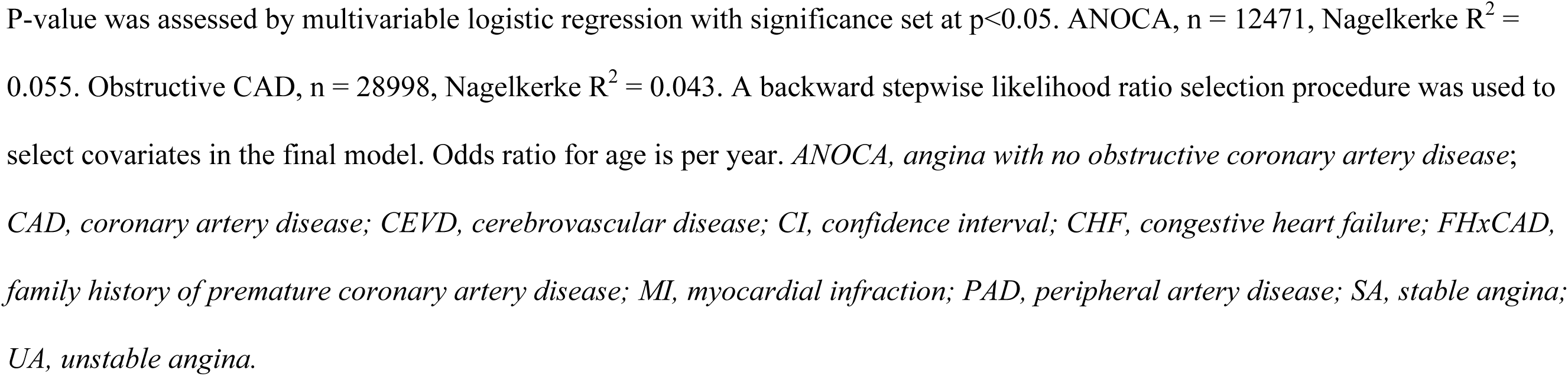
Multivariable logistic regression model defining the factors associated with hospitalizations in ANOCA and obstructive CAD.

| Baseline Characteristics | Odds Ratio | 95% CI for Odds Ratio |  | P-value |
| --- | --- | --- | --- | --- |
|  |  | Lower | Upper |  |
| ANOCA |  |  |  |  |
| Age | 1.03 | 1.02 | 1.03 | <0.001 |
| CEVD | 1.39 | 1.11 | 1.73 | 0.004 |
| CHF | 2.06 | 1.66 | 2.56 | <0.001 |
| Diabetes | 1.24 | 1.10 | 1.40 | <0.001 |
| Hypertension | 1.26 | 1.14 | 1.40 | <0.001 |
| Obesity | 1.15 | 1.04 | 1.27 | 0.006 |
| PAD | 1.89 | 1.43 | 2.50 | <0.001 |
| SA (reference) |  |  |  |  |
| MI vs SA | 1.27 | 1.13 | 1.44 | <0.001 |
| UA vs SA | 1.36 | 1.22 | 1.52 | <0.001 |
| Male vs Female | 1.10 | 1.00 | 1.22 | 0.049 |
| Smoking | 1.16 | 1.06 | 1.28 | 0.002 |
| Obstructive CAD |  |  |  |  |
| Age | 1.01 | 1.01 | 1.01 | <0.001 |
| CEVD | 1.34 | 1.19 | 1.51 | <0.001 |
| CHF | 1.47 | 1.27 | 1.70 | <0.001 |
| Dyslipidemia | 1.17 | 1.10 | 1.24 | <0.001 |
| Diabetes | 1.38 | 1.30 | 1.47 | <0.001 |
| FHxCAD | 1.09 | 1.04 | 1.15 | <0.001 |
| Hypertension | 1.13 | 1.07 | 1.19 | <0.001 |
| Obesity | 1.05 | 1.00 | 1.11 | 0.067 |
| PAD | 1.35 | 1.18 | 1.55 | <0.001 |
| Renal disease | 1.36 | 1.20 | 1.54 | <0.001 |
| SA (reference) |  |  |  |  |
| MI vs SA | 0.67 | 0.63 | 0.71 | <0.001 |
| UA vs SA | 0.86 | 0.80 | 0.92 | <0.001 |

P-value was assessed by multivariable logistic regression with significance set at $p < 0.05$ . ANOCA, $n = 12471$ , Nagelkerke $R^2 = 0.055$ . Obstructive CAD, $n = 28998$ , Nagelkerke $R^2 = 0.043$ . A backward stepwise likelihood ratio selection procedure was used to select covariates in the final model. Odds ratio for age is per year. *ANOCA*, *angina with no obstructive coronary artery disease*; *CAD*, *coronary artery disease*; *CEVD*, *cerebrovascular disease*; *CI*, *confidence interval*; *CHF*, *congestive heart failure*; *FHxCAD*, *family history of premature coronary artery disease*; *MI*, *myocardial infarction*; *PAD*, *peripheral artery disease*; *SA*, *stable angina*; *UA*, *unstable angina*.

## Discussion

To our knowledge, our study is the first to characterize ED visits and assess recent temporal trends of ED visits and hospitalizations in individuals with ANOCA. Notably, we found that while ED visits within two years post-catheterization have remained stable over time, individuals with obstructive CAD experience these visits more frequently than their ANOCA counterparts.

Furthermore, hospitalization within two years post-catheterization was more prevalent among individuals with obstructive CAD but showed a significant decline over time in both groups.

### Temporal trends in ED visits and hospitalizations

Our study detected no changes in ED visits for ANOCA and obstructive CAD but found a significant reduction in hospitalizations for both conditions. This likely resulted from improved outpatient management approaches, including urgent assessment clinics, broader use of risk scores, more accessible access to outpatient investigations, and broader application of observation status.^21–25^ The decline in hospitalizations was particularly notable for obstructive CAD, highlighting the efficacy of robust management strategies frequently used for this demographic.^26^ However, this study also highlighted decreased ED visits among individuals with ANOCA and an MI presentation and decreasing hospitalizations for those with ANOCA and either MI or UA. These findings, while interesting, necessitate further exploration to understand the interventions that are driving these results.

Existing literature in this field provides a context to our findings. Jespersen et al.^7^ assessed rates of cardiovascular hospitalizations and repeat cardiac catheterizations in individuals with SA and no obstructive coronary artery disease (NOCAD). They identified higher rates of these end- points compared to a healthy reference group, with no sex differences in risk of cardiovascular hospitalization after age adjustment. While temporal trends were not assessed by Jespersen et al.,^7^ the hospitalization rates were significantly higher than in the present study. Our study discovered no sex differences in hospitalization rates for individuals diagnosed with ANOCA and either SA or MI. This finding is consistent with a meta-analysis conducted by Chaudhary et al.,^27^ which investigated re-hospitalization rates among individuals with MI and NOCAD. Lastly, Shaw et al.^5^ assessed direct and indirect costs of healthcare resource use among females enrolled in the National Institutes of Health–National Heart, Lung, and Blood Institute–sponsored Women’s Ischemia Syndrome Evaluation (WISE). They reported that females with ANOCA had a 1.8-fold higher angina hospitalization rate than females with 1-vessel CAD and were approximately four times more likely than males to be re-hospitalized for ACS or chest pain. Our study found a declining rate of hospitalizations for individuals with ANOCA, less than those with obstructive CAD, and similar hospitalization rates for males and females. The discrepancies could stem from differences in study design, when the studies were done, and geographical healthcare system differences. Additionally, Shaw et al.’s study was conducted using a highly symptomatic cohort within the WISE study, which could contribute to their findings of a higher rate of angina hospitalization in females with ANOCA.

Our findings also underline the relevance of enhancing the care of ANOCA patients. A clinical trial revealed that individuals with ANOCA, experienced significant impairment in quality of life.^6^ Consequently, each ED visit or hospitalization for such individuals could further deteriorate their quality of life, emphasizing the importance of our findings and the need for further research to develop effective interventions.

### Factors associated with ED visits and hospitalizations

We identified several comorbidities associated with ED visits (Table 3) and hospitalizations (Table 4) among individuals with ANOCA and obstructive CAD. Our findings underscore the complex interplay of comorbid conditions contributing to increased resource use in these populations. CEVD was associated with ED visits and hospitalizations among individuals with ANOCA and obstructive CAD. This is consistent with a prior study demonstrating that cerebrovascular accidents account for many annual cardiovascular hospitalizations.^28^ Similarly, PAD emerged as a significant factor in both groups, aligning with the evidence of PAD accounting for high rates of recurring re-hospitalizations.^29^ CHF was associated with ED visits and hospitalizations among individuals with ANOCA, emphasizing the significant burden that heart failure poses on healthcare utilization in this population. Prior research has similarly found CHF to be a driver of hospital admissions among individuals with CAD.^28, 30, 31^ Interestingly, a history of MI or UA was associated with increased ED visits among those with ANOCA, yet in individuals with obstructive CAD, these factors were associated with a lower odd of hospitalization. This discrepancy might reflect differences in the management of these conditions among individuals with ANOCA and obstructive CAD. We also found hypertension to be associated with hospitalizations among people with ANOCA. Hypertension is a well-established risk factor for cardiovascular disease and has been associated with increased hospitalization rates.^28, 32–34^ In contrast, among those with obstructive CAD, a history of diabetes and renal disease surfaced as factors associated with hospitalization, which is consistent with the extensive literature highlighting the impact of these comorbid conditions on cardiovascular outcomes and hospital admissions.^35, 36^ These findings suggest that individuals with these conditions are at an increased risk of healthcare use, particularly due to their need for frequent follow-ups, management of symptomatic episodes, and treatment of complications.^28^ The identification of these factors emphasizes the importance of comprehensive cardiovascular care in individuals with ANOCA to reduce ED visits and hospitalizations.^28^

### Clinical implications

The stable prevalence of ED visits for ANOCA may reflect a lack of disease management programs and patients’ comfort in self-care or communicating worsening symptoms post-discharge.^37^ The decrease in hospitalizations speaks also to a general decline in the use of in-hospital resources for both groups. Enhanced local education about ANOCA may also be responsible for decreased admissions for this group over time. While ED visits and hospitalizations may not be as frequent as previously suggested, they can still negatively impact quality of life. Given the regular ED attendance of individuals with ANOCA, it’s crucial to educate both patients and care providers about the persistent nature of ANOCA-related chest pain and the potential non-necessity of ED visits. Emerging technologies like the Patient Activation Measure and artificial intelligence could aid in predicting avoidable hospitalizations, allowing physicians to allocate more resources to preventative care.^38, 39^ Additionally, specialized chest pain clinics could provide a multidisciplinary approach to patient care, potentially reducing unnecessary ED visits and enhancing the quality of life for ANOCA patients.^4^

### Limitations

While our study has several strengths, it also shares limitations common to two previous studies.^40, 41^ Our variables were confined to the data elements within data sources, leading to limited or unavailable information on several variables such as presenting symptoms, premature menopause, autoimmune inflammatory diseases, and history of pregnancy-associated conditions.^41, 42^ The presence of individuals with spontaneous coronary artery dissection (SCAD) within the MI cohort could not be ruled out due to the lack of systematic SCAD classification.^43^ A significant number of individuals were excluded due to missing ED visits and hospitalizations data (n=35493), but we anticipate minimal impact on the generalizability of our results given the large remaining sample size (n=85573). The study’s regional focus on Alberta may limit the applicability of referral patterns for non-invasive testing to other regions. The lack of data on non-invasive and invasive ischemia testing made it difficult to comment on ANOCA pathophysiology.

## Conclusions

Our study demonstrated no temporal change in ED visits and a decline in hospitalizations over time for individuals with ANOCA, with no discernible difference between males and females. This trend was also observed in individuals with obstructive CAD, who, contradicting prior studies, showed higher resource use than individuals with ANOCA. Nevertheless, the reduced hospitalizations suggest improved recognition of ANOCA, underlining the importance of ongoing patient education and follow-ups for optimal management.

## Data Availability

The data underlying this study were provided by Alberta Health Services and Alberta Provincial Project for Outcome Assessment in Coronary Heart Disease (APPROACH) under the terms of a research agreement. Inquiries regarding access to the data can be made directly to them.

## Non-standard Abbreviations and Acronyms

ACS: acute coronary syndrome
ANOCA: angina with no obstructive coronary artery disease
APPROACH: Alberta Provincial Project for Outcome Assessment in Coronary Heart Disease
CABG: coronary artery bypass grafting
CAD: coronary artery disease
CEVD: cerebrovascular disease
CIHI: Canadian Institute for Health Information
DAD: Discharge Abstract Database
ED: emergency department
FhxCAD: family history of premature coronary artery disease
LVEF: left ventricular ejection fraction
MI: myocardial infarction
NACRS: National Ambulatory Care Reporting System Database
NSTEMI: non-ST-elevation myocardial infarction
PAD: peripheral artery disease
PCI: percutaneous coronary intervention
SA: stable angina
STEMI: ST-elevation myocardial infarction
UA: unstable angina

## Acknowledgements

We wish to thank the dedicated cardiac catheterization laboratory staff who have entered the data over the years, the SPOR and NACRS team for their help with data extraction, and the Libin Mozell statistical team for helping with data restructuring.

